# Stroke-induced slow oscillations in dynamic brain organization

**DOI:** 10.1101/2024.04.07.24305437

**Authors:** Caroline Tscherpel, Maike Mustin, Nils Rosjat, Ulf Ziemann, Gereon R. Fink, Silvia Daun, Christian Grefkes

## Abstract

A focal ischemic lesion is thought to alter neuronal activity beyond the area of structural damage and thereby interfere with the whole network architecture.

We used a combination of transcranial magnetic stimulation and electroencephalography (TMS-EEG) together with dynamic connectivity analyses and graph theory to study alterations and reorganization of cortical reactivity and connectivity in a cohort of 42 stroke patients longitudinally after stroke. We found a link between an increase of low-frequency coupling in the delta band and alterations in neural information processing in the first weeks after stroke and their relevance for motor recovery more than three months later. We demonstrate that stroke to the motor system leading to impaired hand motor function induce an enhancement of slow activity and delta coupling between frontocentral and parietal regions. In addition, we could show a loss of the physiological network architecture with a decrease of small-worldness and modularity in the delta frequency, implying that a focal ischemic lesion distant to the cerebral cortex interferes with both cortical information integration as well as functional segregation within the days and weeks after stroke. While we found a link between bifrontal coupling in the alpha spectrum and the degree of the motor deficit in the early post-acute phase, the amount of small worldness disruption early after stroke indicated the motor outcome three months later. In contrast, recovery of motor function and cortical reorganization after more than three months post-stroke were paralleled by a normalization of increased low-frequency coupling and a reinstatement of the complex network structure featuring a small-world and modular topology.

## INTRODUCTION

Functional segregation and integration are two contrary yet complementary fundamental organizational principles of the human brain [Tononi et al., 1994]. Functional segregation refers to information processing in distinct, specialized and highly intraconnected subdomains. A segregated neuronal architecture of the brain has been conclusively shown evident at multiple scales in multiple scales in neural networks [Tononi et al., 1998; Wig, 2017]. However, the synchronized processing between the local activity of various functional domains, i.e., functional integration, is equally crucial for effective network function. Hence, the balanced interplay between segregation and integration forms the basis of brain functionality and cognitive abilities [Deco et al., 2015; Wang et al., 2021].

Graph theory has equipped network neuroscience with a language to describe and interpret the complex network organization of the brain [Bertolero and Bassett, 2020; Bullmore and Sporns, 2009]. Implementing this approach, the structural and functional network of the brain could be conceived, unraveling insight into organizational properties and topologies. For both structural and functional brain networks, in human as well as animal models, a ubiquitous configuration in complex systems constitutes the small-world phenomenon [Bassett and Bullmore, 2006; Bullmore and Sporns, 2009; Watts and Strogatz, 1998]. Small-world topology captures a coordinated and balanced pattern of networks with densely intraconnected and sparsely interconnected local modules of nodes. These subnetworks or communities promote functional segregation and yet a minimum path length between distant nodes of distinct subnetworks through long-range connections allowing functional integration. It, furthermore, complies the competitive demands of local and global processing while promoting efficient information flow as well as specialization, adaptability and segregation of information for resilience against focal brain damage [Achard et al., 2006; Bassett and Bullmore, 2006; Bassett and Bullmore, 2017].

Along these lines, stroke is considered as a large-scale network disorder with a focal stroke-induced lesion leading to widespread disruption of interconnected areas distant from the lesion side [Carrera and Tononi, 2014; Grefkes and Fink, 2014]. The concept of diaschisis was coined over a century ago and found revival in the context of connectional diaschisis by advancements in the understanding of evidence of substantial alterations of brain connectivity in the human connectome post-stroke derived by structural [Cheng et al., 2019] and functional MRI [Wang et al., 2010; Yin et al., 2013], leading to changes in network topology in both the ipsi- and contralesional hemisphere. However, studies investigating brain reorganization at the global network level found conflicting results. While some studies suggested that stroke disrupts the balance between local processing and global functioning resulting in a stroke-associated shift towards a more random network architecture [Wang et al., 2010], other findings indicated an increase in global network communication to facilitate information transfer [Blaschke et al., 2021]. A potential resolution to these primarily contradictory results lies in the account for lesion location. Accordingly, damage to those brain regions critical for information processing within a particular subdomain affected functional segregation, while a lesion to a region engaged in communication between subdomains was found to impact on whole brain network organization [Alstott et al., 2009; Honey and Sporns, 2008; Sporns et al., 2007]. What remains elusive, however, is whether alterations of brain networks are a consequence of lesion-induced disconnection and hence may occur in the context of diaschisis or whether they result from plasticity mechanisms supporting recovery and need to be understood in the framework of vicariation.

In this context, it needs to be considered that the fMRI BOLD signal provides only an indirect measure of neuronal activity, reflecting slow-evolving hemodynamic responses not optimally suited to portray the temporal dynamics of neural oscillations [Ozdemir et al., 2020a]. By contrast, EEG directly captures the electrical signal generated by neuronal activity. Furthermore, it also comprehensively maps the entire spectrum of neuronal oscillatory activity which is especially crucial in the framework of stroke, a condition which has been typically shown to be associated with an EEG slowing [Caliandro et al., 2017; Nuwer et al., 1987; Tscherpel et al., 2020]. Although resting-state measures, i.e., either fMRI or EEG, are particularly useful to study stroke patients featuring motor deficits, they only allow an indirect link to function and behavior, the intrinsic resting activity rather yield a weak signal and the underlying activity states are challenging to control. Here, the application of a highly specific, temporally and spatially spatiotemporal dynamics [Deco et al., 2018; Momi et al., 2023]. Combining transcranial magnetic stimulation (TMS) with EEG is a compelling non-invasive perturb-and-measure approach in humans to quantify whole brain cortico-cortical connectivity by probing activity propagation within interconnected networks, inducing synchronization of distant cortical areas and changes in functional dynamics at an individual and millisecond-level [Bortoletto et al., 2015; Momi et al., 2021; Momi et al., 2023]. Particularly relevant in this context, TMS-EEG signal changes follow spatial networks dynamics measured by fMRI, indicating a common neural substrate [Ozdemir et al., 2020b].

Synthesizing the insights above, the main two hypothesis emerges that stroke leads to a large-scale network disruption with alterations of the network topology already in the first days after stroke especially in the low-frequency spectrum. Within the concept of connectional diaschisis these network pathologies normalize with time post-stroke and in parallel to functional recovery.

To this end, we performed graph-theoretic network analyses on dynamic networks based on TMS-EEG connectivity of a cohort of 41 stroke patients with unilateral motor deficits compared to healthy participants. To relate network topology to stroke reorganization and recovery of functional deficits, we analyzed longitudinal TMS-EEG and behavioral data from the first week after stroke until the early chronic stage after more than three months post-stroke.

## MATERIALS AND METHODS

### Participants

Forty-one stroke patients (37 right-handed, 6 females) were recruited and examined at the Department of Neurology, University Hospital of Cologne. Patients were included based on the following inclusion criteria: (i) first ever ischemic stroke with (ii) ≤14 days elapsed from onset and resulting in (iii) unilateral upper limb deficit. Exclusion criteria were (i) contraindication to TMS [Rossi et al., 2021], (ii) bihemispheric infarcts, and (iii) hemorrhagic stroke. In the early chronic phase after more than three months, thirty-five patients underwent a behavioral follow-up visit and twenty-eight patients also participated in a second TMS-EEG assessment (138.3 ± 26.6; 99-183 days). Fifteen age-matched healthy participants without any history of neurological or psychiatric disease (14 right-handed, 2 females) served as a control group. Of note, parts of the original data, i.e., data from 28 patients obtained in the first two weeks after stroke, and the healthy controls’ data, were previously included in [Tscherpel et al., 2020]. Importantly, the current work is motivated and builds up on the finding of slow activity after stroke presented in the former publication. Nevertheless, there is no overlap in the analyses as the former did not incorporate any connectivity analyses. Thus, all analyses presented here are entirely novel. All participants provided informed written consent before inclusion into the study. The ethics committee of the Medical faculty of at the University of Cologne ethically approved the study (file no. 17-244) which was carried out following the Declaration of Helsinki (October 2013).

The following motor and clinical scores were assessed within (i) the first two weeks and (ii) more than three months post-stroke: Stroke severity and the global neurological impairment were assessed by the NIHSS (http://www.ninds.nih.gov/doctors/NIH_Stroke_Scale.pdf), the Action Research Arm Test (ARAT) allowed to evaluate gross and fine upper limb functions including pinch, grasp and grip [Lyle, 1981], the Motricity Index (MI) classifying muscular strength in the proximal, middle, and distal joints of arms and legs [Demeurisse et al., 1980], and maximum grip strength as determined by a vigorimeter (KLS Martin Group, Germany).

### TMS parameters

A focal 70mm figure-of-eight alpha coil connected to a Magstim Super Rapid^2^ stimulator (The Magstim Co. Ltd, Whitland, United Kingdom) was used to perform transcranial magnetic stimulation (TMS). To test the integrity and functionality of the motor network after stroke, we used the ipsilesional primary motor cortex (M1) as the stimulation target. M1 was defined by the motor hotspot, i.e., the position of the TMS coil held tangentially to the skull in a 45° posterior-anterior direction eliciting motor evoked potentials (MEP) of the highest amplitudes of the first interosseous (FDI) muscle. Ag-AgCl self-adhesive electrodes were placed over the muscle according to the belly-tendon montage. The EMG signal was amplified, high- and band-pass filtered (0.5Hz high-pass; 30-300Hz band-pass), and digitized using a Powerlab 26T device combined with the LabChart software (Package version 8.0; AD Instruments, Sydney, Australia).

TMS was controlled by a frameless computerized stereotactic neuronavigation system throughout the experiment (BrainSight V.2.0.7; Rogue Research Ltd; Montreal, Canada). For this purpose, the head of the participants was co-registered with an individual anatomical MR image which allowed to reliably monitor online the position of the TMS coil in reference to the stimulation coordinated and the subject’s head, within and across sessions.

The resting motor threshold (RMT) was individually defined using TMS Motor Threshold Assessment Tool (MTAT) 2.0 (http://www.clinicalresearcher.org/software.html) [Awiszus, 2003]. In twenty-three patients we could not determine a MEP in response to the stimulation of ipsilesional M1, even with maximal stimulator output. Here, the RMT of contralesional M1 served as a reference for the individual threshold [Tscherpel et al., 2020]. Additionally, in these cases the ipsilesional M1 was anatomically identified as the hand knob on the precentral gyrus.

### EEG recordings

TMS-EEG was recorded using a 64-channel TMS-compatible EEG system (BrainAmp DC, BrainProducts GmbH, Gilching, Germany) that prevents amplifier saturation and reduces the induced magnetic artefacts. The EEG signals were sampled at 5kHz with a resolution of 0.1µV per bit, and high-pass filtered mounted on an elastic electrode cap (EasyCap) following the international 10-20 system. Horizontal and vertical eye movements were recorded using the two remaining electrodes as electrooculogram (EOG) sensors. Impedances of all electrodes were kept below 5kΩ throughout the whole experiment. Participants wore inserted earplugs to prevent EEG auditory evoked potentials induced by the TMS click, p [Braack et al., 2015; Ilmoniemi and Kičić, 2009; Nikouline et al., 1999]. Masking the TMS click with white noise [Belardinelli et al., 2019] have proven difficult in acute stroke patients. Using the former approach, we have previously shown to capture a biologically relevant neural signal largely without major confounds of peripheral co-activation [Tscherpel et al., 2020]. Moreover, bone conduction produced by TMS was attenuated by placing a thin layer of film between the coil and EEG cap [Massimini et al., 2005; Massimini et al., 2007; Tscherpel et al., 2020].

### Experimental procedure

During the EEG recordings, subjects were seated in a comfortable chair with eyes open. Severely affected stroke patients, who were unable to sit due to their deficit, were assessed in their beds with a 45° incline of the headboard (n=23).

For each TMS-EEG session we collected at least 100 trials of single pulses with an interstimulus interval randomly jittered between 6.5-8.0s with 80% RMT. This intensity range is considered to be above the threshold for a significant EEG response, but still minimize re-afferent somatosensory feedback impacting on the EEG reponse [Casali et al., 2010; Fecchio et al., 2017; Komssi et al., 2004; Rosanova et al., 2009; Tscherpel et al., 2020].

## Data analysis

### Preprocessing

Data analysis was performed using Matlab (The MathWorks, Massachusetts, USA). Artifacts contaminated channels and trials were visually inspected and manually rejected [Casarotto et al., 2016; Rosanova et al., 2018; Tscherpel et al., 2020]. Recordings with either less than 90 artifact-free trials or to TMS pulse onset was removed and missing data was replaced with baseline [Rogasch et al., 2016; Tscherpel et al., 2020]. Subsequently, EEG data were detrended, band-pass and band-stop filtered (1-60Hz; 49-51Hz; Butterworth 3^rd^ order), down-sampled at 625Hz, and segmented in time windows of ±1000ms around the TMS pulse [Casarotto et al., 2016; Tscherpel et al., 2020]. Bad channels were interpolated using EEGLAB spherical interpolation function (https://sccn.ucsd.edu/eeglab/) [Delorme and Makeig, 2004]. Signals were average re-referenced and baseline corrected [Casarotto et al., 2016]. Finally, independent component analysis (ICA, EEGLAB runica function) was applied to remove residual ocular, muscle, or TMS-related artifacts [Rogasch et al., 2014].

For all analyses, data from patients with right-hemispheric lesions (n=23) were flipped along the midsagittal plane so that the lesioned side and thus the stimulation site corresponded to the left hemisphere in all participants. Correspondingly, data from healthy participants, in which the right hemisphere was tested, also were flipped to account for systematic effects from hemispheric differences.

### Construction of phase-locking networks

Before connectivity analysis, EEG data was spatially filtered using surface Laplacian, which is considered to substantially reduce the effect of volume conduction by enhancing localized activity and suppressing diffused signals and, thus, allows the analysis of electrodes close to the region of interest [Lachaux et al., 1999; Rosjat et al., 2021]. Therefore, preprocessed epochs were re-referenced to the Laplacian reference (http://psychophysiology.cpmc.columbia.edu/Software/CSDtoolbox) to improve spatial resolution and the suitability for subsequent connectivity analysis [Hjorth, 1975; Kayser and Tenke, 2006]. Of note, edge electrodes were excluded from analyses, leading to a total of 39 electrodes for further investigation.

Subsequently, data was decomposed to the time-frequency domain using Morlet wavelet transform (5 cycles) [KRONLAND-MARTINET et al., 1987] allowing to compute the temporal evolution of amplitude and phase for each frequency separately in the delta- to beta-frequency range (1-30Hz). This procedure Human Neuroimaging, London, UK, http://www.fil.ion.ucl.ac.uk) implemented in Matlab. Amplitude and phase information were further analyzed using customized Matlab scripts.

We quantified the connection between two brain regions by the synchronization between the activity of two sites as determined by the single-frequency phase-locking value at the sensor level (PLV; adapting the phase-locking value defined in [Lachaux et al., 1999]) [Rosjat et al., 2018; Rosjat et al., 2021], resulting in a positive number between 0 and 1 with PLV=1 occuring in cases of perfect intertrial phase locking of the phase difference between the EEG signals of a pair of two channels over all trials. We, here, considered the four frequency bands: δ (1-4Hz), θ (4-8Hz), α (8-12Hz) and β (13-30Hz). We calculated the PLV separately for each frequency with a resolution of 1Hz and subsequently averaged the values over the respective frequency band [Rosjat et al., 2021]. This analysis yielded 760 connectivity values for each frequency and each participant.

To test, whether changes averaged over frequency bands relative to pre-stimulus baseline differ significantly either between stroke patients and healthy controls or the acute and chronic phase, we compared PLV values obtained for each pair of electrodes at each time point of the post-stimulus interval of interest, i.e., 0-600ms, between groups. To this end, we used a pointwise t-test with a significance level of p<0.05 FDR-corrected for multiple comparisons. The corrections were performed concerning the number of time points, groups, and electrodes [Rosjat et al., 2021].

### Applying graph theoretical measures

After the construction of the PLV networks for each frequency band, undirected graphs with nodes formed by sensors and edges representing PLV values, were constructed and analyzed using graph theoretical metrics as implemented in the Brain Connectivity Toolbox (BCT) (http://www.brain-connectivity-toolbox.net) [Rubinov and Sporns, 2010]. Based on previous publications applying graph theory on networks after stroke, we used a threshold of 0.5 to convert connectivity matrices into weighted networks, which means that PLV values higher than the applied threshold were referred to as edge weights [Wang et al., 2010]. Notably in order to account for time-varying connectivity patterns, occur [Sizemore and Bassett, 2018]. In addition, we also analyzed static networks which constitute aggregated graphs that consider the number of occurrences of each edge.

We confined our analyses predominantly to parameters characterizing the whole network. However, based on the finding of increased ipsilesional frontoparietal connectivity after stroke, we next analyzed connections of the ipsi- and contralesional network separately using only sensors on the left or right hemisphere as well as edges within the centroparietal (FC1-FC3-C1-C3-CP3-P1-P3/FC2-FC4-C2-C4-CP4-P2-P4) or frontal network (Fz-F1-F3-AF3/Fz-F2-F4-AF4). In detail, we assessed (i) node strength as the sum of all weighted edges of connected nodes indexing a parameter of network connectivity and connectivity strength, (ii) the network core which is the largest subnetwork comprising all nodes with degree of at least two which identifies the most tightly connected parts of a graph providing insights into the overall structure of a networks [Rubinov and Sporns, 2010], (iii) small worldness as the ratio of the normalized mean clustering coefficient (CC) and normalized characteristic path length (CPL). The clustering coefficient calculates the proportion of existing clusters to all possible clusters. A cluster is defined as a triangle of connections between two connected neighbors of one node. The mean clustering coefficients, reflecting the degree of functional specialization, is normalized to the average mean clustering coefficient of 100 random networks with the same number of nodes, edges, and degree distribution [Blaschke et al., 2021; Rubinov and Sporns, 2010]. The shortest path represents the smallest number of edges that connect two nodes. Here, we used the mean average of shortest path length between all pairs of nodes in the network, normalized to a random network as described above. Complementary to CC, the CPL is used as a measure of integration within the network of interest. Crucially, small worldness as the ratio of CC and CPL with high values of CC and small values of CPL enables both segregated and specialized but also distributed and integrated information processing. Thus, small world typology is suggested to constitute effective information processing structure, minimizing wiring cost while offering dynamical complexity [Bassett and Bullmore, 2006; Bassett and Bullmore, 2016]. Finally, we calculated (iv) modularity by Newman’s spectral community detection algorithm [Rubinov and Sporns, 2010], quantifying how much a network can be subdivided into minimizes the number of between-modular edges. Consequently, modularity is supposed to represent the amount of functional specificity and information segregation within a network [Bertolero et al., 2015].

### Statistical analysis

Statistical analyses were performed using the software SPSS (Statistical Package for the Social Science, version 28, IBM). Group differences between stroke patients and healthy individuals were evaluated by either independent two-sided t-test (p<0.05). In case of dynamic measures with multiple time periods, we performed repeated measures analyses of variance (ANOVA) (5 level: 0-50; 50-100; 100-200; 200-400; 400-600) x frequency (4 level: DELTA; THETA; ALPHA; BETA) X group (2 level: patients; controls)). Furthermore, Pearson correlations were computed to test for linear associations between behavioral scores and graph parameters. The behavioral scores were used as a motor composite score generated by a principal component analysis [Tscherpel et al., 2020]. Post-hoc tests and correlation analyses were FDR-corrected for multiple comparisons.

## RESULTS

### Increased ipsilesional parietal integration in the early phase after stroke

Analyzing the TMS-evoked connectivity, i.e. the phase-locking value of TMS-EEG signals, aggregated over time revealed that, compared to healthy control subjects, stroke patients in the first week after stroke featured an increase in coupling between frontocentral and parietal areas in the ipsilesional hemisphere particularly in the low-frequency delta and theta, but also in the alpha range (p<0.05, FDR-corrected, Figure 1A). Additionally, stroke patients showed enhanced coupling in response to the TMS perturbation in primary motor areas of both the ipsilesional and the contralesional hemisphere in the aforementioned spectra. Of note, for the beta frequency, stroke patients exhibited reduced connections in the ipsilesional hemisphere, especially between central and frontal regions (p<0.05, FDR-corrected, Figure 1A).

**Figure 1.**
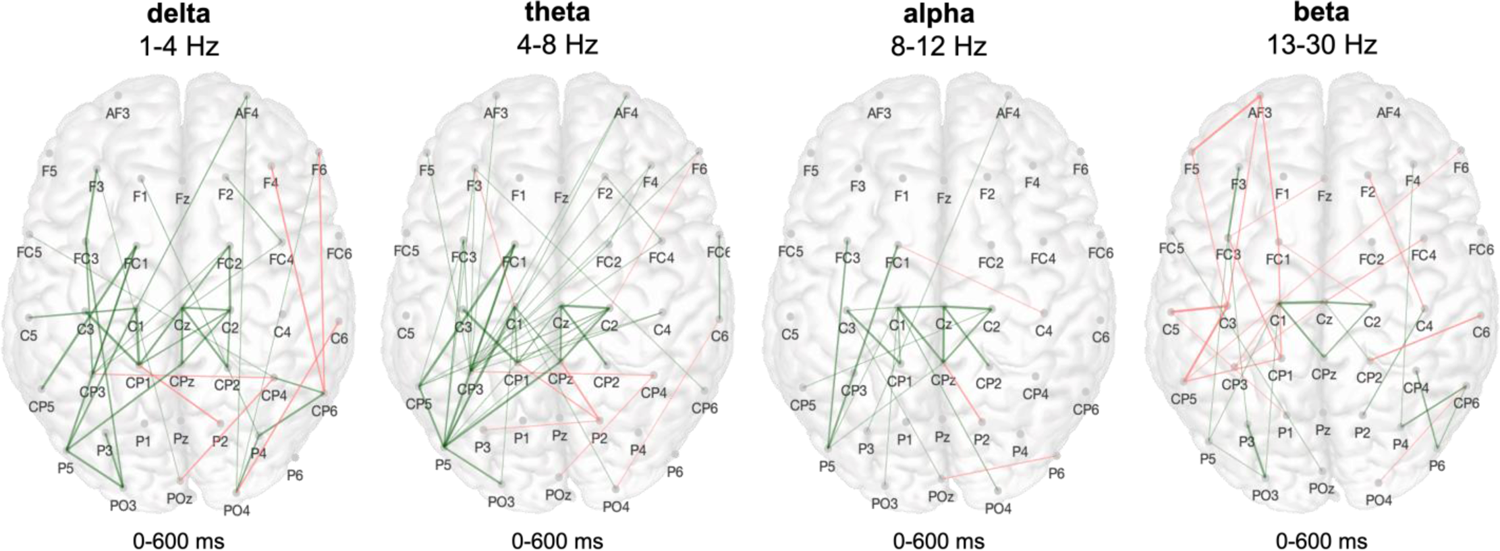
Increase of low frequency coupling in frontoparietal areas early after stroke Aggregated network connections over time. Please note, that connections that appear more often are displayed with bolder graphs between sensors. The between-group comparison between healthy controls and stroke patients early after stroke is depicted illustrating the increase of slow oscillation coupling early after stroke (green: stroke>healthy participants; red: stroke<healthy participants).

To further quantify the alterations of the network architecture post-stroke, also in a temporal context, we next employed graph-theoretical analyses. Assessing the connection strength between all nodes in the network, analyses of the whole post-stimulus phase indicated an exclusive increase in node strength for the delta band early after stroke (delta: p=0.03, t_(50.4)_=2.70; theta: p=0.18, t_(54.0)_=1.71; alpha: p=0.36, t_(54.0)_=1.11; beta: p=0.46, t_(54.0)_=0.75; FDR-corrected). However, mapping the time course of node strength dynamically over the time after the TMS pulse (Figure 2) suggested differential curve characteristics. Thus, considering distinctive time periods after TMS onset, we found significant between-group differences for node strength depending on the frequency band as well as on time post-stimulus, as revealed by a significant interaction effect for TIME X FREQUENCY X GROUP (F(12,648)=2.04, p=0.02) (main effect TIME: F(4,216)=11.28, p<0.001; main effect: FREQUENCY F(3,162)=25.12, p<0.001; interaction effect TIME X GROUP: F_(4,216)_=2.78, p=0.028; interaction effect FREQUENCY X GROUP: F_(3,162)_=2.78, p=0.096). Post-hoc test revealed that early after stroke node strength was increased for all frequency bands within the first 50ms after TMS onset early after stroke (delta: p=0.006, t_(53.4)_=3.16; theta: p=0.32, t_(54.0)_=2.20; alpha: p=0.007, t_(52.0)_=2.82; beta: p=0.006, t_(53.8)_=3.24; FDR-corrected). In the delta and theta range, the between-group difference also lasted throughout the time period between 50ms and 100ms (delta: p=0.014, t_(53.8)_=3.03; theta: p=0.014, t_(33.4)_=2.93; alpha: p=0.07, t_(52.1)_=1.81; beta: was only significant in the delta band (delta: p=0.024, t_(54.0)_=2.84; theta: p=0.38, t_(54.0)_=1.33; alpha: p=0.94, t_(54.0)_=0.07; beta: p=0.94, t_(54.0)_=0.13; FDR-corrected) and remained solely enhanced here until 400ms post-stimulus (delta: p=0.028, t_(51.3)_=2.54; theta: p=0.51, t_(54.0)_=0.88; alpha: p=0.72, t_(54.0)_=0.37; beta: p=0.72, t_(54.0)_=-0.37; FDR-corrected; 400ms-600ms: all p-values≥0.4).

**Figure 2.**
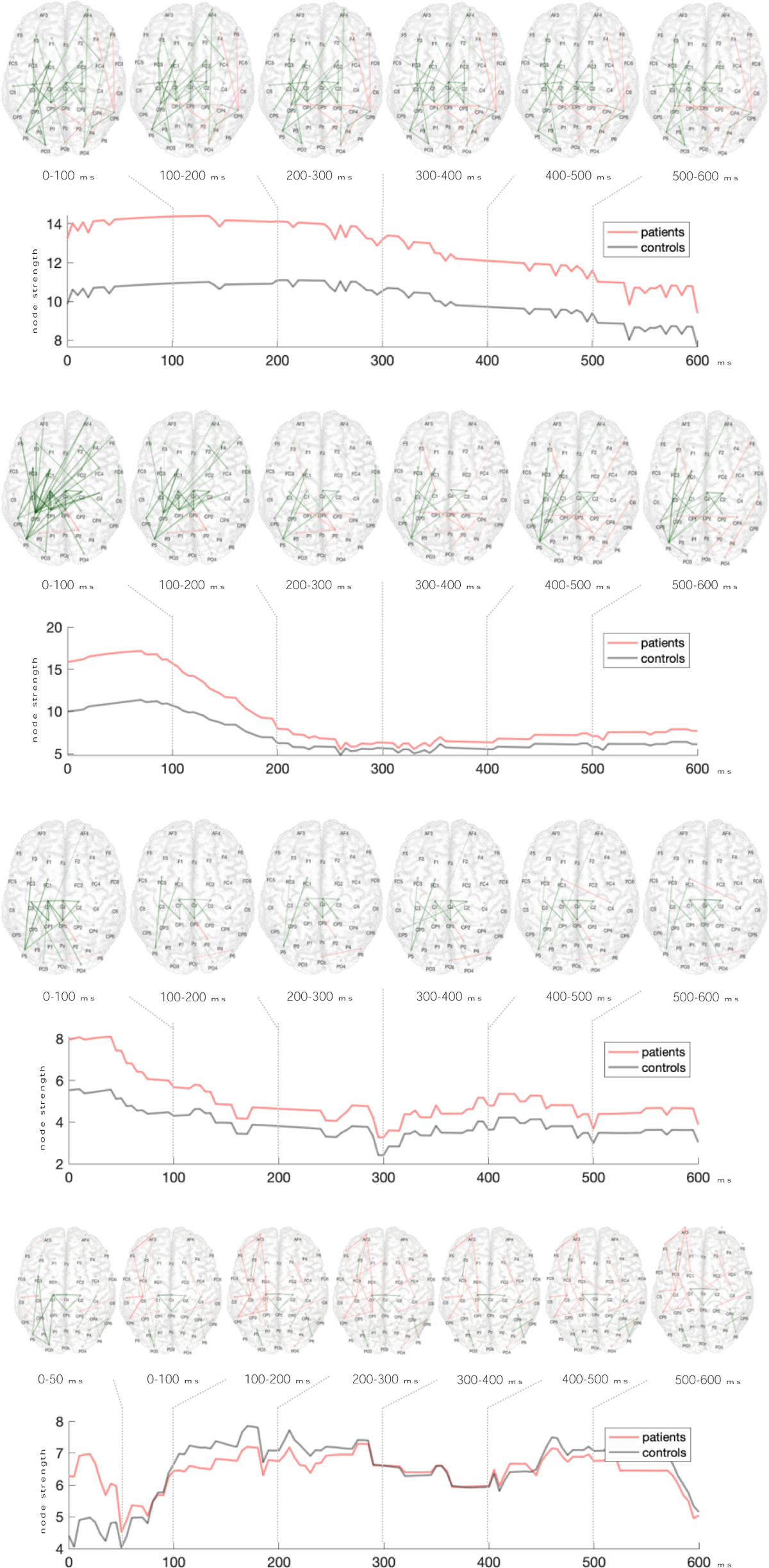
Dynamic connectivity early post-stroke for each frequency band The upper panel displays the dynamic network connections along the post-stimulus period aggregated over 100 ms (colors indicate between group comparison of connectivity with green: stroke>healthy participants and red: stroke<healthy participants) and the lower panel shows the time course of overall node strength (red: patients, grey: control subjects).

In consequence, we focused on the delta and theta frequency band for subsequent graph theoretical analyses. As implied by the differences in connectivity matrices (Fig. 1A), the increase in node strength early after stroke was predominantly driven by coupling effects in the ipsilesional hemisphere (delta: p=0.012, t_(53.0)_=2.77; theta: p=0.012, t_(45.6)_=2.62; FDR-corrected), and here mainly by edges between frontocentral and parietal nodes (delta: p<0.001, t_(53.6)_=3.92; theta: p<0.001, t_(51.9)_=4.44; FDR-corrected). Notably, neither node strength of the contralesional hemisphere nor of ipsilesional frontal nodes showed significant differences between patients early after stroke and healthy participants (contralateral nodes: delta: p=0.16, t_(54.0)_=2.08; theta: p=0.16, t_(54.0)_=1.77; ipsilesional frontal nodes: delta: p=0.79, t_(54.0)_=0.27; theta: p=0.53, t_(54.0)_=0.85), underlining the region-specificity of the findings.

### Extensive network post-stroke with reduced efficiency of information flow

Complementary, global network properties were also altered early after stroke. TMS-evoked interregional coupling exhibited a spatially broader network core in the low-frequency range of delta and theta (delta: p=0.011, t_(54.0)_=2.77; theta: p=0.011, t_(16.9)_=2.85; FDR-corrected), I.e., the largest subnetwork comprising all nodes with degree of at least two [Rubinov and Sporns, 2010]. Portraying the significant core nodes over the time post-stimulus illustrated an extensive bihemispheric network topology that lasted up to 600 ms post-stimulus (Figure 3). While in stroke patients TMS-induced coupling formed a core comprising bihemispheric centroparietal and ipsilesional parietal nodes over the entire post-stimulus period, especially in the delta frequency band, in healthy controls the core nodes recruited by TMS predominantly involved frontal and frontocentral nodes.

**Figure 3.**
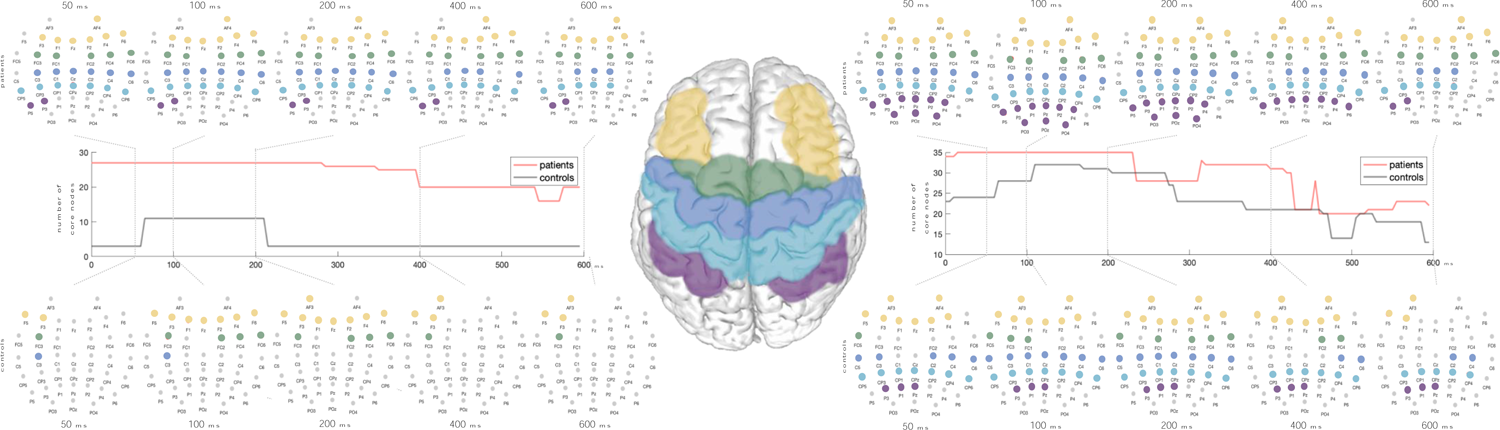
Stroke is associated to a broader network core comprising bihemispheric centroparietal and ipsilesional parietal nodes The significant core nodes over the time post-stimulus are illustrated with an extensive bihemispheric network topology that lasted up to 600 ms post-stimulus in the delta and theta-frequency in stroke patients. Upper panel displays the network core of stroke patients, in the middle the time course of significant core nodes for the entire post-stimulus period and the lower panel shows the network of healthy controls.

However, the architecture of networks can not only be characterized by the involved nodes and connection strength inbetween. The complex network organization of the brain hold properties of information for resilience. Accordingly, in the first weeks after stroke the network evoked by TMS featured a substantial decrease in small worldness in the delta range (delta: p=0.016, t_(54.0)_=-2.74; theta: p=0.62, t_(54.0)_=0.31; FDR-corrected). Crucially, the decrease in small worldness was predominantly caused by an increase in path length (p=0.009, t_(50.4)_=2.70). Thus, early after stroke, signal propagation from specialized brain regions require considerably more processing steps for the integration of information, and thus substantially higher costs.

In addition, the network alterations in the first weeks after stroke could be also characterized by a deterioration of modularity for the delta frequency (delta: p=0.038, t_(30.8)_=-2.17; theta: p=0.17 t_(54.0)_=-1.38; FDR-corrected), implying a reduction of organization in functionally specific subnetworks and thereby a decrease in information segregation early after stroke.

Notably, while alterations of node strength were mainly attributed to the ipsilesional hemisphere, stroke-associated changes of the higher network structure were found in both, i.e., the ipsilesional and the contralesional hemisphere (small worldness: ipsilateral: p<0.001, t_(54.0)_=-3.54, contralateral: p=0.031, t_(54.0)_=-2.22; modularity: ipsilateral: p=0.025, t_(34.8)_=-2.52, contralateral: p=0.025, t_(54.0)_=-2.43; FDR-corrected).

### Bifrontal connections underlying acute motor function post-stroke

Despite the profound alterations of the network configuration in the first weeks after stroke, there was no association between the observed changes in the low-frequency bands and the clinical deficit assessed in the early phase after stroke, i.e., NIHSS and motor scores (all p-values>0.2). Thus, the parietal shift in TMS-induced coupling and the reduction of a modular and small-world network topology the delta frequency rather seems to be a general phenomenon not specifically linked to the acute clinical deficit.

To further explore network dynamics underlying motor function after stroke, we divided the patient group featuring a broad range of clinical deficits (grip strength: 28.2 ± 35.7; 0-94.7; ARAT: 18.1 ± 22.7; 0-55; MI: 42.5 ± 34.0; 0-95.5; NIHSS: 7.5 ± 3.5; 1-14) into two subgroups using k-means cluster analysis represented a subgroup of severely affected patients (n=25; grip strength: 2.0 ± 6.9; 0-29.3; ARAT: 0.9 ± 1.5; 0-3; MI: 18.1 ± 15.8; 0-58; NIHSS: 9.8 ± 1.9; 1-14) and a subgroup representing patients with mild to moderate motor deficits (n=16; grip strength: 69.2 ± 19.9; 43.5-94.7; ARAT: 44.9 ± 10.2; 27-55; MI: 80.5 ± 12.7; 47-95.5; NIHSS: 3.8 ± 2.1; 1-8).

Contrasting the TMS-evoked coupling between the both subgroups of patients, i.e. severely affected patients and moderately to mildly affect patients, revealed that severely affected patients featured a reduction of contralesional frontal connections particularly in the alpha frequency but also in theta and beta range (p<0.05, FDR-corrected, Figure 4A). Likewise, in the alpha band, decreased frontal coupling could be also found in the ipsilesional hemisphere.

**Figure 4.**
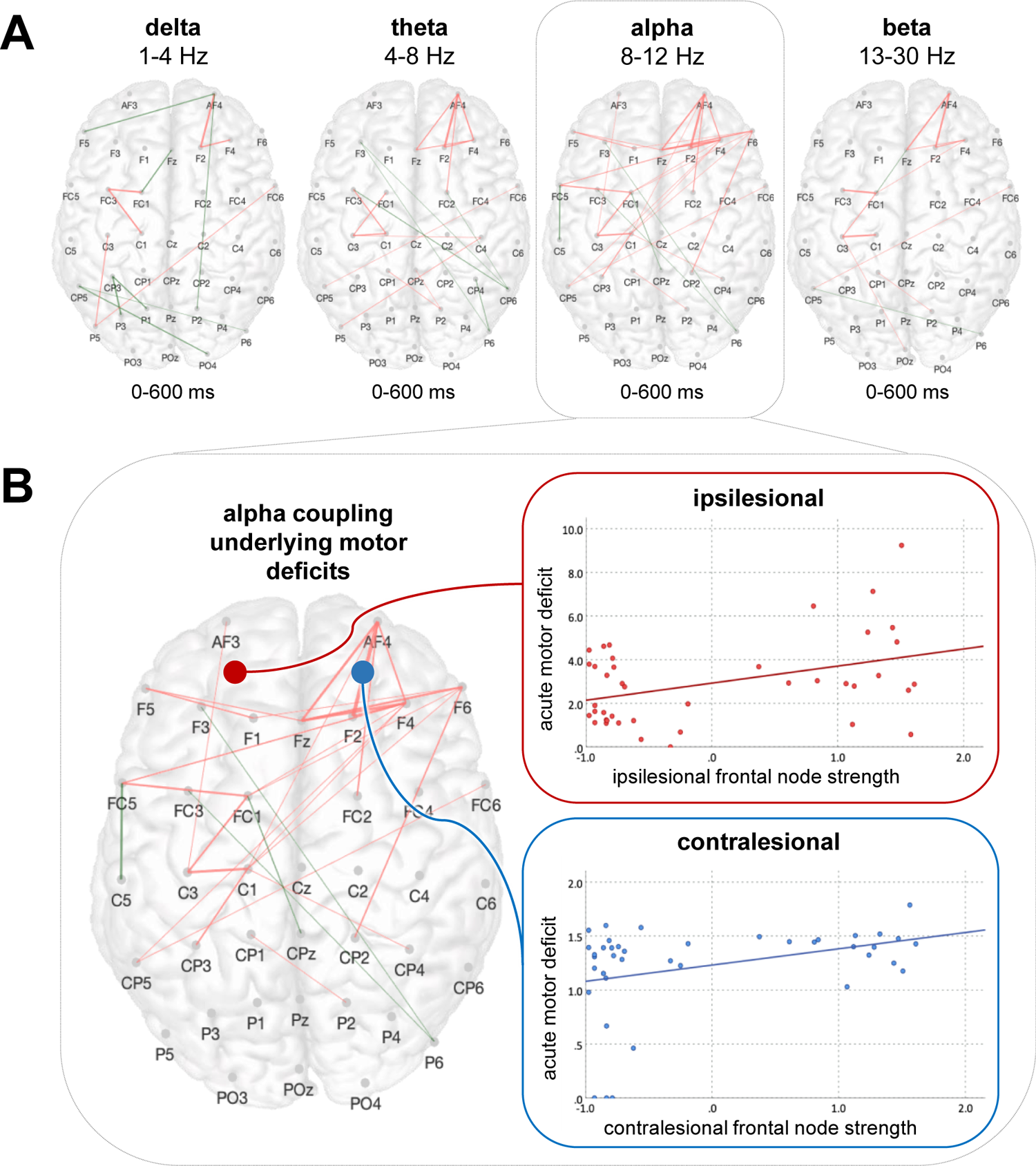
Bifrontal connections are indicative of the motor deficit early after stroke (A) Between-group comparisons between the subgroups of severely affected patients and mildly to moderately affected patients indicated a decrease of node strength in more affected patients in the alpha band in ipsilesional as well as contralesional frontal areas. (B) Alpha node strength of frontal connections in both, i.e., the ipsilesional and the contralesional hemisphere was linked to the acute motor deficit.

Correspondingly, between-group comparisons indicated a decrease of node strength in severely affected patients compared to mildly to moderately impaired patients in the alpha band in ipsilesional as well as contralesional frontal areas (ipsilesional: delta: p=0.22, t_(39.0)_=-1.36; theta: p=0.09, t_(39.0)_=-2.09; alpha: p=0.016, t_(39.0)_=-3.09; beta: p=0.22, t_(22.1)_=-1.26; contralesional: delta: p=0.75, t_(39.0)_=-0.32; theta: p=0.53, t_(39.0)_=-0.91; alpha: p=0.04, t_(32.5)_=-2.74; beta: p=0.53, t_(39.0)_=-0.85; FDR-corrected). Beyond that, alpha node strength of frontal connections in both, i.e., the ipsilesional and the contralesional hemisphere was associated with the composite score of the acute motor deficit (ipsilesional: r=0.41, p=0.016; contralesional: r=0.35, p=0.025; FDR-corrected), indicating that stronger TMS-evoked bifrontal alpha coupling underlies better motor function early after stroke (Figure 4B).

### Reorganization of TMS-evoked coupling and recovery post-stroke

After more than three months post-stroke, stroke patients experienced substantial motor recovery (p<0.001, t_(34.0)_=-4.80) (Figure 5B). In parallel, the connectivity map of reorganization, i.e., the significant group differences in coupling aggregated over time post-stimulus between the three months follow up and the first weeks post stroke, showed a decrease of connections particularly between ipsilesional frontocentral and parietal areas in the low-frequency range of delta, theta, and alpha (p<0.05, FDR-corrected, Figure 5A).

**Figure 5.**
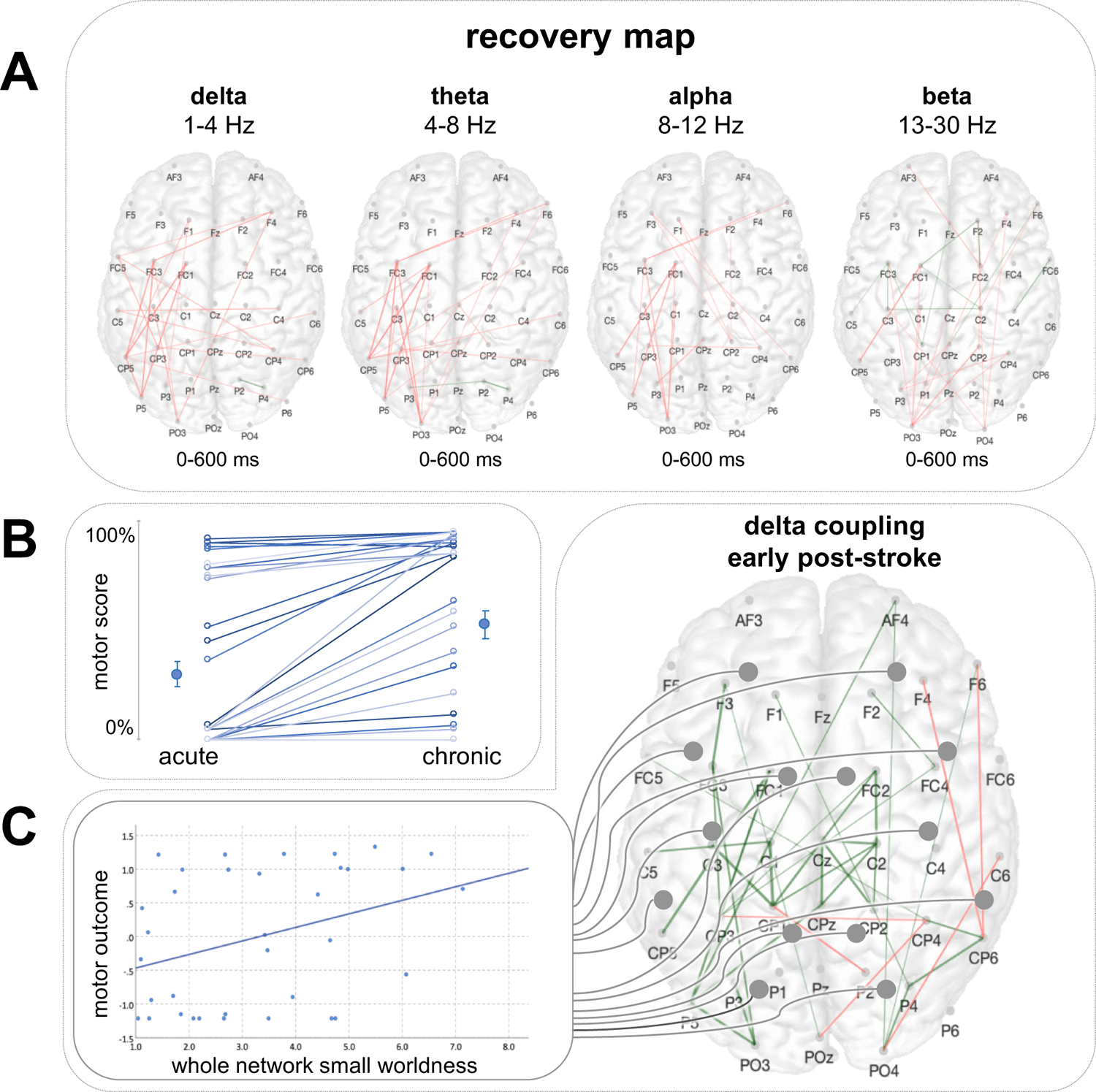
Motor recovery after stroke is paralleled by a normalization of increased delta coupling (A) Aggregated network connections over time of within-group comparison of stroke patients early after stroke and in the chronic post-stroke phase (red: acute>chronic). (B) Motor recovery and motor reorganization is paralleled by a decrease of initially augmented coupling in parietofrontal areas in the low frequency range. (C) By contrast, the decrease of small worldness early post-stroke in the delta range is indicative of a poor motor outcome.

Here, node strength of frontoparietal coupling in the ipsilesional hemisphere, which was increased in comparison to healthy participants in the first weeks post-stroke in the delta and theta frequency, significantly decreased in these frequency bands and also in the alpha band (delta: p=0.034, t_(65.9)_=1.86; theta: p=0.026, t_(67.0)_=2.30; alpha: p=0.026, t_(67.0)_=2.16;, FDR-corrected) during motor reorganization. Along with this normalization of the initially enhanced interregional coupling in low-frequency spectra, characteristics of the complex network structure further strive to reinstate. Correspondingly, the reduction of small worldness in the delta frequency found early post-stroke again improved after more than three months after stroke (p=0.029, t_(55.8)_=-1.93). Likewise, we found a significant increase of modularity in the delta frequency at follow-up three months later (p=0.036, t_(66.7)_=-1.83). Thus, properties of complex network organization which we showed to be substantially impaired early after stroke demonstrate an amelioration in parallel to motor recovery.

Finally and crucially, the acute decrease in small worldness, early after stroke was linked to the motor outcome after more than three months post-stroke (r=0.37, p=0.029). Thus, patients featuring a particular decrease in small world topology in the delta band showed a less favorable motor outcome (Figure 5C).

Combining TMS-EEG, dynamic connectivity analyses and graph theory we uniquely draw a link between an increase of low-frequency coupling in the delta band and alterations of the complex network topology early after stroke to their functional relevance for motor recovery in the chronic post-stroke phase. Specifically, we demonstrate that stroke is not only associated with an increase of slow activity but that stroke lesions also induce an enhancement of delta coupling between frontocentral and parietal regions, replicating the finding of parietal involvement of fMRI studies. However, at the level of the higher network structure, we also found a loss of the physiological network architecture with a decrease of small-worldness and modularity in the delta frequency, implying a stroke-associated alteration of both information integration as well as segregation within the first weeks after stroke. While bifrontal connections in the alpha spectrum seem to be most crucial for the functional deficit in the early post-acute phase, solely the impairment of small-worldness in the delta frequency was indicative of a more pronounced permanent deficit. In contrast, recovery of motor function and cortical reorganization after more than six months post-stroke were paralleled a normalization of increased low-frequency coupling and a reinstatement of the complex network structure featuring a small-world and modular topology.

### Connectivity, randomness and diaschisis after stroke

In line with our TMS-EEG data, the finding of stroke-induced changes in functional and effective connectivity of the intra- and interhemispheric network architecture has been substantiated by fMRI studies (see e.g. [Grefkes and Fink, 2014] for review). Accordingly, reorganization of the motor network after stroke encompasses not only an acute reduction of interhemispheric connections as well as connectivity between ipsilesional M1 and frontoparietal regions very early after stroke, but also a gradual increase and restoration of network interactions in the following weeks and months [Golestani et al., 2013; Meer et al., 2010; Park et al., 2011]. Furthermore, neural networks reorganizing post-stroke showed increased faciliatory coupling between M1 and fronto-parietal areas in the affected hemisphere [Rehme et al., 2011; Rehme and Grefkes, 2013]. In particular, most analyses have focused on frontal from fMRI studies for increased connectivity to ipsilesional parietal cortex in stroke patients, paralleling the present results [Backhaus et al., 2021; Park et al., 2011; Schulz et al., 2016]. As the reconstitution of physiological connectivity patterns have been associated with a favorable recovery in the chronic phase, these network alterations are suggested to be functionally relevant [Grefkes and Fink, 2014; Rehme et al., 2011]. However, the body of fMRI connectivity studies have still not elucidated if changes of connectivity enable functional recovery or merely represent the neuronal aftermaths of adaptation and compensation. For parietofrontal connections, both enhanced fMRI coupling within the first weeks post-stroke and increased connectivity strength in the chronic phase were associated to a less favorable chronic outcome and, thus, a more severe permanent impairment respectively [Backhaus et al., 2021; Schulz et al., 2016]. This upregulation has been interpreted as a manifestation of compensation and additional recruitment of parietal areas involved in sensory processing and sensorimotor integration [Reibelt et al., 2023]. Although we found a comparable enhancement of coupling between frontocentral and parietal regions, the unique methodological combination of TMS-EEG, connectivity and graph theory will however dispute the previous understanding. In particular, by linking increased frontoparietal coupling early after stroke to the loss of the physiological network architecture, our data rather question the compensatory nature.

Accordingly, beyond investigating changes in specific functional subnetworks and away from descriptive anatomy towards measures of network efficiency and functionality, the application of graph theory on brain connectivity data allows to disclose the lesion’s impact on the global network organization and its complex relationship within and between communities of highly connected nodes promoting functional segregation while preserving functional integration, thus, potentially reflecting the complexity of behavioral deficits more accurately [Bassett and Bullmore, 2006; Sporns, 2013]. The small-world organization is particularly important for the resilience of the brain architecture and for protecting the integrity of its networks [Achard et al., 2006]. In an experimental stroke model, an increase of small-worldness of functional connectivity in the somatosensory network was found in the subacute phase – suggesting to reflect excessive neuronal re-wiring - which shifted to baseline topology with recovery of topology is underpinned by a translational approach with a comparable pattern of network parameters in the early subacute phase in mice and humans, indicating a facilitation of the global communication structure [Blaschke et al., 2021]. The alterations’ functional significance was rather heterogeneous: While the increase in small-worldness in the group of mildly-affected patients was predominantly found in case of greater motor impairment, the initial increase of small-world topology was related to a more favorable outcome in the chronic phase in the mouse model [Blaschke et al., 2021].

In contrast, other fMRI studies have reported a decrease of small-world topology after stroke [Siegel et al., 2018; Wang et al., 2010] which underlines our present findings. Accordingly, in a longitudinal study of subcortical strokes leading to severe deficits, the motor network gradually shifted towards a random mode with time after stroke during the recovery process and in association to impairment, suggesting that a less optimized network reorganization is involved in regaining motor function [Wang et al., 2010]. Furthermore, and similarly to the present results, Siegel and colleagues found a decrease of small-worldness and modularity already in the early post-acute phase [Siegel et al., 2018], implying that both integration within and segregation between the functional networks are reduced early after stroke. Again in line with our results, the return of a modular network featuring a small-world topology partially recovered over time and in parallel to functional recovery [Siegel et al., 2018]. Complementary evidences also stem from EEG studies which have shown a frequency-dependent modulation of the small world properties with an increase of small-worldness in the alpha band but a stroke-associated decrease in the delta frequency [Caliandro et al., 2017].

Of note, a shift towards network randomization was also shown for other neuropathologies as brain tumor, severe traumatic brain injury or Alzheimer’s disease, hypothetically implying a potential common pathway of the brain’ reaction to a lesion or neurodegenerative processes [Bartolomei et al., 2006; Gerloff and Hallett, 2010; Nakamura et al., 2009; Stam et al., 2007]. Although the underlying substrates for network randomization remain speculative, potential explanations might lie in a compensatory but non-optimized outgrowth of new connections because of structural or functional disconnection of physiological pathways, modulation of synaptic activity in the networks, alterations of the timing of and Hallett, 2010; Meer et al., 2010; Newbold and Dosenbach, 2021]. Along these lines, alterations of the network dynamic, more precisely, a shift towards randomization have to be considered also as a manifestation of diaschisis, naturalistically revealing a non-optimized architecture. To be recognized as diaschisis, i.e., alterations ought to be most pronounced early after stroke, progressively normalize with time and in parallel to function and are related to behavior [Carrera and Tononi, 2014]. Whether these alterations of the network structure after stroke, however, may arise either in the context of diaschisis or from plasticity mechanisms beneficial for functional recovery after stroke have yet remained unsolved.

Here, a causative factor might constitute the fact that most of the former studies are based on either resting-state or structural connectivity, only allowing an indirect link to function and the specific network modality. Although there is an obvious advantage for resting measures in patients with neurological deficits, a selective activation of specific networks might be necessary to delineate its precise functional relevance [Gerloff and Hallett, 2010]. Moreover, the BOLD signal constitutes an indirect measure of neuronal activity with relatively limited temporal resolution, likely not optimally suited to capture rapidly varying, subtle neuronal processes. By contrast, EEG and MEG directly record faster oscillatory signals and phase information in and across frequencies of rhythmic oscillations which have been shown to be profoundly relevant for sensorimotor and cognitive functions [Llinás et al., 1999] and linked to GABAergic as well as glutamatergic activity [Premoli et al., 2014; Premoli et al., 2017]. Thus, by providing information about oscillatory properties, both modalities add an additional functional layer to connectivity, unavailable for fMRI techniques [Bortoletto et al., 2015].

However, the spontaneous resting EEG is a compound of different activity states of distinct areas and thus challenging to control and steady over time, even under standardized conditions [Rosanova et al., 2009]. Within this context, the direct activation of a specific region by means of an intense and selective perturbation such as TMS offers a systematic and causal measure of the region’s intrinsic properties and connections [Rosanova et al., 2009]. In consequence, the methodology of TMS-EEG not only allows an unrivaled activation of the motor system as well as the assessment of effective connectivity by network but also to elucidate pathophysiological mechanisms underlying alterations of connectivity through the incorporation of oscillatory information.

Accordingly, at the level of anatomic subnetworks, we demonstrated an increased coupling within parieto-frontal circuits early after stroke which subsequently and progressively decrease with time post-stroke. At the level of the global network architecture, we found a decrease in modularity and small-worldness in the first weeks after stroke, followed by a restoration of the physiological network architecture after more than three months post-stroke along with functional improvement. In the dimension of oscillations, these alterations were predominantly evident for slow rhythms particularly in the delta frequency. Crucially, the decrease in small-worldness in the delta band was associated to the functional outcome in the chronic stroke phase with initially more pronounced alterations of the network architecture leading to a less favorable outcome. In summary, the observed stroke-induced connectivity changes in the slow frequency range need to be understood in the context of diaschisis as defined above, implying a link between diaschisis, network randomization, the familiar phenomenon of a stroke-associated EEG slowing, and the functional implications of delta activity. Hence, a profound understanding of our results regarding the network architecture after stroke demands to extensively comprehend the findings of stroke-related changes of neuronal oscillations.

Linking increased slow activity early after stroke to network randomization and post-stroke recovery An increase of slow activity particularly in the perilesional vicinity but also in areas distant from the lesion within the unaffected hemisphere has been described as the predominant finding in early EEG recordings post-stroke almost half a century ago [Gloor et al., 1977; Nuwer et al., 1987]. Ever since, elevated activity of slow oscillations, characteristically quantified as low-frequency power, has been detected in both animal models [Carmichael and Chesselet, 2002; Facchin et al., 2020; Nita et al., 2007] and patient studies, across electrophysiological methodologies [Tecchio et al., 2006] as well as the timespan from the acute to the chronic stroke phase [Finnigan and Putten, 2013; Wu et al., 2016]. In the ipsilesional hemisphere, augmented delta activity has often been found to be accompanied with a Finnigan et al., 2007; Wijngaarden et al., 2016]. From a clinical perspective, alone or in combination with an ipsilesional alpha power decrease, ipsi- and contralesional EEG slowing have been associated with a poor outcome post-stroke [Finnigan and Putten, 2013]. More mechanistically, delta waves were shown to be related to stroke-induced ischemia and tissue dysfunction [Ferreira et al., 2021].

However, although the underlying mechanisms of physiological slow activity have been distinctly identified, the neural underpinnings of stroke-associated slow oscillations remain unclarified. Notably in this context, the terms of slow activity, indicating the faster spectrum of delta power, and sleep-like slow waves, referring to slow oscillatory activity <1Hz as the manifestation of off-periods and cortical bistability [Compte et al., 2003; Steriade et al., 1993; Timofeev et al., 2001], are not always unambiguously separable in the literature, although recent work has demonstrated different regulation principles and, thus, potentially distinct underlying mechanistic circuits [Hubbard et al., 2020]. While the occurrence of sleep-like slow waves in stroke patients [Sarasso et al., 2020; Tscherpel et al., 2020] has also been shown lately, the present work is rather signified to slow activity in the delta range from 1-4Hz.

Delta waves are an EEG hallmark of physiological NREM sleep generated and modulated by the thalamocortical circuitry [Uygun and Basheer, 2022]. Accordingly, thalamocortical relay neurons project to the cortex and - in an hyperpolarized voltage range - hold an intrinsic burst-firing activity, constituting the generator of cortical delta waves [McCormick and Pape, 1990; Steriade et al., 1991]. This self-oscillatory activity arises from the orchestrated interplay between hyperpolarization-activated cyclic nucleotide-gated (HCN) channels and T-type voltage gated calcium (CaV) channels as well as rapidly deactivating voltage sensitive potassium (KV) channels [Cain and Snutch, 2013; Choi et al., 2015; McCormick and Pape, 1990; Rudy and McBain, 2001]. However, since thalamocortical neurons only generate delta waves in hyperpolarization and burst-firing mode, inhibitory inputs are crucial for the initialization and promotion of delta waves, resulting from inhibitory GABAergic input of the thalamic reticular nucleus [Lewis et al., 2015; Uygun and Basheer, 2022]. In contrast, glutamatergic excitatory of thalamocortical neurons and, thus, inhibit delta [Brown et al., 2012; Franks, 2008].

Crucially in this context, HCN as well as CaV channels have been shown to be involved in the pathological process of ischemic brain injury [Chen et al., 2020; Dai et al., 2023]. Moreover, there is evidence for increased tonic inhibition governed by extra-synaptic GABAA receptors after stroke [Clarkson et al., 2010]. As pharmacological studies revealed that under physiological conditions enhancing the GABAergic influence on thalamocortical neurons mediated through extra-synaptic GABAA receptors enhances delta waves [Cope et al., 2005; Mesbah-Oskui et al., 2014; Uygun et al., 2022], similar neuronal mechanisms determined for NREM delta waves may equally apply to the underpinnings of increased slow activity after stroke. This appears particularly relevant in view of the contemporary understanding that sleep does not necessarily occur exclusively at the behavioral level as a distinct global state, but also during wakefulness locally confined to discrete cortical areas with detrimental behavioral consequences [Nir et al., 2011; Sarasso et al., 2020; Vyazovskiy et al., 2011], again opening a parallel to slow activity after stroke and stroke-induced functional deficits.

Likewise, the deceleration of alpha power post-stroke [Dubovik et al., 2012; Finnigan et al., 2007; Wijngaarden et al., 2016] accompanied with the increase of slow activity is also well explained within the thalamocortical circuitry.

Accordingly, alpha rhythms are strongly influenced by the thalamus and underlie the same cellular components of thalamocortical relay neurons outlined above, albeit under depolarized conditions for example when activated by cortical glutamatergic input [Hughes et al., 2004; Hughes and Crunelli, 2016]. In other words, alterations of thalamocortical network dynamics associated with attenuated neocortical drive onto the thalamus, i.e., deafferentation and, thus, thalamic disfacilitation and pathologically high levels of inhibition as found in stroke lead to hyperpolarization and, thereby, to both the depression of alpha activity and increase of slower frequencies [Hughes and Crunelli, 2016; Wijngaarden et al., 2016].

Notably, as argued above, we understand the increase of frontoparietal coupling as well as the alteration of the functional network architecture, i.e., the network randomization, early after stroke in we link connectional diaschisis after stroke to the disruption of thalamocortical circuits. In other ways, pathological lesion-induced thalamo-cortical dynamics propagate low-frequency oscillations beyond the lesion through thalamocortical pathways in widespread areas of the neocortex [Wijngaarden et al., 2016]. The resulting connections form an overconnected, random network that lack a resilient and optimized small-world topology.

In accordance with our findings, inactivation of a cortical node by chemogenetic inhibition led to increased connectivity between the inhibited area and its direct thalamocortical target in an animal model [Rocchi et al., 2022]. Using in vivo electrophysiology, the authors could also show an enhancement of low frequency oscillations via the suppression of neural firing, resulting in increased delta coherence between regions that exhibited increased coupling [Rocchi et al., 2022].

It was suggested quite early that disruptions within thalamocortical circuits with excessive expression of low-frequency bursting in thalamic nuclei and their large-scale propagation through the neocortex result in neurological symptoms in several conditions including motor symptoms in Parkinson’s disease [Llinás et al., 1999]. Accordingly, deep brain stimulation targeting thalamic nuclei and their afferent structures is supposed to reinstate neurophysiological function and modulate pathologically altered circuits [Lozano and Lipsman, 2013]. Although this notion is less prevalent in stroke studies, several references imply a similarly substantial disturbance of thalamocortical network dynamics after stroke. For example, the thalamus and thalamocortical connections in the affected hemisphere have been shown to play a crucial role for neuronal reorganization and functional recovery after stroke [Markus et al., 2005]. Furthermore, using EEG and a computational model it has been demonstrated that stroke lesions lead to excessive hyperpolarization of thalamic neurons, that switch into burst-firing mode and this, in turn, entrains thalamo-cortical pathways, propagating low-frequency oscillations into widespread areas of the neocortex [Wijngaarden et al., 2016]. Moreover, optogenetic stimulation of ipsilesional M1 in a stroke mice model restored activation in the ipsilesional corticothalamic circuit and improved functional recovery, underlining the involvement of corticothalamic loops in recovery after stroke [Vahdat et al., 2021].

One causative factor leading to the disruption of thalamocortical circuits that need to be considered particularly in the context of stroke is the notion of disconnection. First, stroke-induced lesions do not only affect cortical output, but also interrupt input fibers and thus yield to a cortical deafferentation and decreased modulations of ascending activating systems [Grefkes and Fink, 2014]. This is supported by findings from TMS studies indicating that corticospinal tract lesions activate inhibitory circuits [Classen et al., 1997; Liepert et al., 2005]. On the other hand, lesions are associated with an attenuated neocortical drive onto the thalamus [Wijngaarden et al., 2016]. Along these lines, a critical reduction of cortico-cortical connectivity has also been suggested to alter the balance of inhibition and excitation [Funk et al., 2017; Mann et al., 2009]. Experimental recordings from deafferented cortical slabs isolated from thalamic and cortical inputs have revealed an amplification of slow oscillations further underlining the link between disconnection and slow activity [Timofeev et al., 2000].

Finally, the critical question of the functional implications of increased delta connectivity and network randomization for motor reorganization after stroke still remains to be answered: Delta waves as found in physiological conditions seem to be critically implicated in many processes that benefit memory, cognition, and health, including synaptic homeostasis, cellular energy regulation, clearance of toxic proteins, and neuronal plasticity [Dworak et al., 2010; Tononi and Cirelli, 2006; Tononi and Cirelli, 2014; Xie et al., 2013]. While, theoretically, our findings might be understood as a representation of recovery processes similar to physiological sleep and increased connectivity in the framework of a compensatory re-wiring, our data rather imply an association to an attenuated recovery processes and persistent disability in the chronic phase after stroke. In other words, low-frequency activity after stroke hinders the dynamic brain organization, in turn leading to poor functional outcome. In contrast, motor recovery was paralleled by a restoration of the complex network organization, indicating that reorganization after stroke signifies a return to information integration for efficiency and information segregation for resilience.

## CONCLUSION

We here identified increased frontoparietal low-frequency coupling and its deterioration of a physiological network architecture early after stroke in the context of diaschisis. By adding the functional layer of oscillational properties to connectivity, we revise the understanding of the neural substrate of diaschisis as well as remote effect of focal lesions and associate them with a disturbance of thalamocortical circuits. This notion also indicates to rethink treatment strategies of post-stroke deficits and neuromodulatory interventions, particularly suggesting subcortical structures as a target for therapy and demanding techniques capable to reach these deeper regions [Baker et al., 2023; Wessel et al., 2023].

## Data Availability

All data produced in the present study are available upon reasonable request to the authors

## ACKNOWLEDGMENTS

GRF and CG are funded by the Deutsche Forschungsgemeinschaft (DFG, German Research Foundation) – Project-ID 431549029 – SFB 1451 (project C05). We are grateful to Sebastian Dern, Jana Freytag, Sebastian Günther, Natascha Kellner, and Julien Schuckelt for their technical assistance.

